# Development and Cross-Database External Validation of 24-Hour Landmark Models for ICU Mortality After Intracerebral Hemorrhage: Incremental Value of Routine Coagulation Markers

**DOI:** 10.64898/2026.10.11.26365237

**Authors:** Sakshie Pathak, Yong Si, Janet Sanjaya, Mohammadsaeed Haghi, Nausin Tabassum Kudrot, Greg Placencia, Kamiar Alaei, Maryam Pishgar

## Abstract

**Background:** Intracerebral hemorrhage (ICH) is associated with high early mortality. Although routine coagulation markers are clinically relevant, their incremental value for mortality prediction beyond neurologic and physiologic information is uncertain. We developed and externally validated models using first-24-hour ICU data and evaluated the added predictive value of routine coagulation measurements.

**Methods:** We used MIMIC-IV for model development and eICU-CRD for external validation. The development cohort included 2,669 adults with nontraumatic ICH and 388 ICU deaths. Logistic regression, random forest, and XGBoost were evaluated using a patient-level 70:30 holdout split and 5-fold stratified cross-validation. External validation used a high-specificity primary eICU cohort (396 patients; 39 deaths) and a broader secondary hemorrhagic-stroke/likely-ICH cohort (1,555 patients; 168 deaths). Performance was assessed using AUROC, AUPRC, Brier score, calibration, and paired bootstrap comparisons. GCS-only and age-plus-GCS models served as clinical benchmarks. Coagulation-only, clinical-only, and combined XGBoost models were compared to assess incremental value.

**Results:** In the internal holdout cohort, AUROCs were 0.870 for logistic regression, 0.859 for random forest, and 0.867 for XGBoost. In the primary external cohort, corresponding AUROCs were 0.804, 0.839, and 0.817; AUPRCs were 0.342, 0.419, and 0.406; and Brier scores were 0.080, 0.072, and 0.074. In the broader cohort, AUROCs were 0.826, 0.848, and 0.838. Random forest showed the numerically strongest external performance, but pairwise AUROC differences among multivariable models were not clearly supported. GCS-only achieved AUROCs of 0.780 and 0.813 in the primary and broader cohorts. Coagulation-only XGBoost showed weak discrimination (AUROC 0.598 internally, 0.598 primary external, and 0.566 broader external). Adding coagulation variables to the clinical model produced AUROC differences of 0.014 (95% CI ***−***0.009 to 0.037) and 0.003 (95% CI ***−***0.007 to 0.013) in the two external cohorts.

**Conclusions:** Multivariable models based on first-24-hour ICU data retained useful discrimination across external cohorts and provided modest gains over simple GCS-based benchmarks. Routine coagulation markers alone were weak predictors and did not provide clearly supported incremental discrimination beyond broader clinical information. Cross-database transportability should be interpreted together with phenotype alignment and calibration.

## 1 Background

Intracerebral hemorrhage (ICH) is among the most severe forms of stroke and remains associated with high early mortality and substantial long-term disability despite advances in acute neurologic and critical care [1–4]. Patients with ICH frequently require intensive care for neurologic monitoring, airway protection, blood-pressure management, and treatment of systemic complications [1, 5]. Early prognostic assessment may therefore support risk stratification, monitoring intensity, communication with patients and families, and the design of clinical decision-support strategies. However, prognostic models intended for this setting must distinguish information available at the intended prediction time from events occurring later during the ICU stay and must remain reliable when applied outside the population in which they were developed.

The ICH Score remains the best-established bedside prognostic tool for spontaneous ICH and combines Glasgow Coma Scale (GCS), age, hematoma volume, intraventricular hemorrhage, and infratentorial location [6]. Its prognostic utility has been demonstrated in external populations [7], and GCS itself remains a strong marker of neurologic severity. Nevertheless, several components of the ICH Score depend on imaging-derived anatomic information that is not consistently represented as structured variables in large electronic health record databases. Models based on routinely available ICU data may therefore provide a complementary approach when the objective is structured, reproducible risk estimation from electronic clinical information rather than replacement of established bedside assessment.

Machine learning has increasingly been applied to clinical risk prediction across critical care, neurologic, and cardiovascular populations [8, 9]. In patients with cerebral hemorrhage or spontaneous ICH, several studies have demonstrated that routinely collected clinical information can support mortality prediction beyond conventional univariable risk assessment. Nie et al. applied multiple machine-learning algorithms to critically ill patients with cerebral hemorrhage and showed that multivariable models could achieve useful discrimination for mortality prediction [10]. More recently, Yap et al. developed machine-learning models specifically for mortality prediction among ICU patients with spontaneous ICH and reported strong predictive performance, with ensemble tree-based methods among the leading approaches [11]. Matsumoto et al. incorporated both clinical and imaging information and compared machine-learning models with the established ICH Score, demonstrating the potential value of richer multimodal predictors for in-hospital mortality estimation [12].

Recent work has also placed greater emphasis on temporal and external validation. Yang et al. developed interpretable models using first-day clinical information from MIMIC-IV and evaluated them in an independent MIMIC-III cohort for 30-day mortality prediction, providing evidence that early structured electronic health record data can retain prognostic value across database versions [13]. Chen et al. extended this concept to cross-database validation by developing time-to-event models in eICU and externally evaluating them in MIMIC-IV [14]. Collectively, these studies establish that machine-learning approaches can support ICH prognostication and that model transport beyond the development sample is feasible. However, important questions remain regarding performance under tightly aligned disease phenotypes, robustness when phenotype definitions are broadened, calibration across databases, and the incremental value of specific routinely measured laboratory domains beyond established neurologic and physiologic predictors.

Coagulation measurements represent a clinically plausible but incompletely characterized component of ICH risk prediction. Platelet count, international normalized ratio (INR), and activated partial thromboplastin time (aPTT) may reflect impaired hemostasis, anticoagulant exposure, or broader systemic illness. Coagulation-derived measures have been associated with hospital outcomes after spontaneous ICH [15]. However, prognostic association does not necessarily imply incremental predictive value. Neurologic severity, age, hemodynamic instability, metabolic abnormalities, and other routinely collected laboratory measurements may already capture much of the risk information associated with adverse outcomes. A clinically relevant modeling question is therefore whether routine coagulation markers contribute additional predictive information once these broader features are incorporated.

Independent external validation is particularly important for this question because prediction performance can change when models are transported across clinical environments. Differences in disease coding, case mix, missing-data patterns, measurement frequency, and variable semantics may affect both discrimination and calibration even when the underlying modeling algorithm is unchanged [16–18]. This issue is especially relevant when combining large critical care databases. MIMIC-IV provides detailed longitudinal data from a single academic medical center, whereas the eICU Collaborative Research Database represents a multicenter US critical care population [19, 20]. At the same time, the two databases encode diagnoses differently: MIMIC-IV provides structured diagnosis codes, whereas eICU relies heavily on hierarchical and free-text diagnosis fields. External validation of an ICH model therefore requires not only transport of the fitted prediction pipeline but also careful alignment of the target phenotype.

In the present study, we developed ICU mortality prediction models using information available during the first 24 hours of ICU care among adults with nontraumatic ICH in MIMIC-IV and externally evaluated the locked modeling pipeline in eICU. To address the tradeoff between phenotype specificity and statistical precision, external validation was performed in two cohorts whose definitions were fixed before final model evaluation: a high-specificity primary cohort designed to align closely with the MIMIC-IV ICH phenotype and a larger secondary hemorrhagic-stroke/likely-ICH cohort used to assess broader transportability. We compared logistic regression, random forest, and XGBoost with GCS-only and age-plus-GCS benchmarks; evaluated discrimination, calibration, and clinical utility; and directly compared coagulationonly, clinical-only, and combined predictor sets. We hypothesized that multivariable first-24-hour models would retain prognostic discrimination across databases, whereas routine coagulation markers would provide limited incremental predictive value once broader neurologic and physiologic information was available.

## 2 Methods

### 2.1 Study Design, Data Sources, and Cohort Definition

We conducted a retrospective cross-database prediction study using the Medical Information Mart for Intensive Care IV database (MIMIC-IV, version 3.1) for model development and internal validation and the eICU Collaborative Research Database (eICU-CRD, version 2.0) for external validation. Both databases contain de-identified electronic health record data from critically ill patients and are available to credentialed researchers through PhysioNet [21, 22]; their database design and characteristics have been described previously [19, 20]. MIMIC-IV contains detailed clinical data from a single academic medical center, whereas eICU-CRD represents a multicenter US critical care population. Reporting was guided by the TRIPOD+AI recommendations for clinical prediction models [18].

The prediction landmark was defined at 24 hours after ICU admission. Predictor information was restricted to the interval from ICU admission through the first 1,440 minutes, and patients were required to remain in the ICU beyond this landmark. The outcome was death between the 24-hour landmark and discharge from the indexed ICU stay. Only the first eligible ICU stay for each patient was retained.

The MIMIC-IV development cohort included adults aged 18 years or older with nontraumatic intracerebral hemorrhage identified using ICD-9 code 431 or ICD-10 code I61 in any hospital diagnosis position. Patients whose ICU stay did not extend beyond 24 hours were excluded. The final development cohort comprised 2,669 unique patients, including 388 ICU deaths.

External validation applied the same age restriction, 24-hour predictor window, ICU length-of-stay requirement, first-eligible-stay rule, and ICU mortality endpoint. Because eICU diagnoses are represented primarily through hierarchical and free-text fields rather than ICD billing codes, two operational external phenotypes were used. Their definitions were fixed before final external model evaluation and were applied unchanged thereafter.

The primary strict cohort used a high-specificity phenotype based on the eICU diagnosis string field, with qualifying terminology permitted in any diagnosis row. Patients were included if diagnosis text contained explicit intracerebral, intraparenchymal, or parenchymal hemorrhage, hematoma, or bleed terminology; a recognized brain location term together with explicit hemorrhage-related terminology; or hemorrhagic stroke/hemorrhagic CVA terminology accompanied by hypertensive or intracerebral/intraparenchymal context. Trauma, subarachnoid hemorrhage, subdural hemorrhage or hematoma, epidural hemorrhage or hematoma, aneurysmal hemorrhage, postoperative or post-craniotomy hemorrhage, CNS mass or tumor-related diagnoses, skull fracture or concussion, and hemorrhagic transformation of ischemic stroke were excluded. Anatomical location terms alone were insufficient for inclusion without hemorrhage-related context. The final strict cohort contained 396 patients and 39 ICU deaths.

The secondary broader cohort was designed to assess transportability under a less restrictive hemorrhagic-stroke/likely-ICH phenotype. It retained all patients meeting the strict phenotype as well as additional broader-eligible stays with explicit hemorrhage-related terminology and no competing exclusion diagnosis, while applying the same adult restriction, ICU length-of-stay threshold, first-24-hour feature window, ICU mortality endpoint, and first-eligible-stay-per-patient rule. The final broader cohort contained 1,555 patients and 168 ICU deaths. At the patient level, all 396 patients in the strict cohort were represented in the broader cohort. Because the first eligible stay was selected separately under each phenotype definition, 393 patients contributed the same indexed ICU stay in both analyses, whereas three contributed an earlier broader-eligible stay in the secondary cohort. Detailed operational inclusion and exclusion criteria for both eICU phenotypes are provided in Additional file 1 (Supplementary Table S1).

In eICU, preserved event offsets were used to verify that predictor measurements occurred within the intended 0–1,440-minute window and that included patients remained in the ICU beyond the prediction landmark. ICU mortality was defined using indexed ICU discharge status, with Expired classified as death.

Both source databases contain de-identified data released under their respective institutional approvals and data-use agreements. This study involved secondary analysis of de-identified data with no patient contact or access to directly identifying information. Database access was obtained after completion of the required credentialing and research training.

### 2.2 Predictors and Preprocessing

The common predictor set was restricted to variables available during the first 24 hours of ICU care and that could be harmonized across MIMIC-IV and eICU-CRD. Predictors included:

- demographic characteristics: age and sex;
- neurologic severity: minimum Glasgow Coma Scale (GCS) score;
- coagulation measurements: minimum platelet count, maximum international normalized ratio (INR), and maximum activated partial thromboplastin time (aPTT);
- vital signs: minimum and maximum heart rate, minimum and maximum respiratory rate, minimum systolic blood pressure, and minimum and maximum temperature; and
- additional laboratory measurements: maximum white blood cell count, maximum blood urea nitrogen, minimum and maximum potassium, minimum and maximum sodium, maximum bicarbonate, and maximum bilirubin.

For repeatedly measured variables, the specified minimum or maximum value observed during the first 24 hours was used. Source-specific variable definitions and measurement units were harmonized before cross-database model application. In eICU, temperature measurements were integrated from vitalPeriodic and nurseCharting, while systolic blood pressure measurements were integrated across vitalPeriodic, vitalAperiodic, and nurseCharting. Temperature values were harmonized to degrees Celsius before first-24-hour summaries were generated.

No complete-case restriction was applied. In the primary analysis, missing predictor values were handled using mean imputation, followed by z-score standardization within a common modeling pipeline. For internal holdout evaluation, imputation and scaling parameters were estimated using the MIMIC-IV training partition only and subsequently applied unchanged to the holdout partition.

For external validation, preprocessing parameters were re-estimated using the complete patient-level MIMIC-IV development cohort after internal evaluation had been completed. The fitted transformation was then applied unchanged to both eICU cohorts. No imputation parameters, scaling parameters, model coefficients, or hyperparameters were estimated using external-validation data.

### 2.3 Model Development and Internal Validation

Three multivariable models were evaluated: logistic regression, random forest, and XGBoost. These approaches were selected to represent a conventional linear probabilistic model and two nonlinear ensemble methods [23, 24].

Logistic regression used L2 regularization with *C* = 1.0, the lbfgs solver, an intercept term, and a maximum of 3,000 iterations. Random forest used 300 trees, Gini impurity, a maximum depth of 10, square-root feature sampling, a minimum of two observations to split an internal node, a minimum leaf size of one, bootstrap sampling, and random state 42. XGBoost used a binary logistic objective with logloss evaluation, 300 boosting rounds, maximum tree depth of 3, learning rate 0.05, subsampling fraction 0.8, column subsampling fraction 0.8, and random state 42. The primary models were trained without class weighting. Model specifications were not modified according to final external-validation performance.

The MIMIC-IV cohort was divided at the patient level using a 70:30 outcome-stratified training–holdout split with random state 42. All preprocessing operations for this analysis were fitted exclusively in the training partition. Five-fold stratified cross-validation was additionally performed to assess the stability of model discrimination, with preprocessing re-estimated within each training fold.

Two parsimonious logistic-regression models were evaluated as clinical benchmarks: a GCS-only model and an age-plus-GCS model. Both used the same logistic-regression specification and preprocessing framework as the primary logistic-regression model. GCS was selected because neurologic severity is a central component of conventional ICH prognostication [6, 25]. A complete conventional ICH Score could not be reconstructed because structured hematoma volume, intraventricular extension, and sufficiently detailed hemorrhage-location information were not consistently available in the harmonized feature set.

After internal evaluation was completed, each model and its preprocessing pipeline were refitted using the complete MIMIC-IV development cohort for subsequent external validation.

### 2.4 External Validation and Performance Evaluation

The refitted MIMIC-IV models were applied directly to the strict primary and broader secondary eICU cohorts without external model updating, hyperparameter optimization, or recalibration.

Discrimination was assessed using the area under the receiver operating characteristic curve (AUROC) and area under the precision-recall curve (AUPRC). Overall probability accuracy was evaluated using the Brier score [16, 26]. External calibration was characterized using calibration intercept, calibration slope, mean predicted risk, observed event rate, and graphical calibration curves [17, 27].

Ninety-five percent confidence intervals for AUROC and AUPRC were estimated using 2,000 patient-level percentile bootstrap resamples. Pairwise differences in AUROC were evaluated using paired bootstrap resampling so that competing models were compared within the same resampled patients. Comparisons were performed among the three multivariable models and between the GCS-only benchmark and each multivariable model. Because the strict external cohort contained only 39 outcome events, its calibration estimates and paired model differences were interpreted cautiously.

### 2.5 Secondary and Sensitivity Analyses

The incremental contribution of routine coagulation measurements was evaluated using three XGBoost predictor sets. The coagulation-only model included minimum platelet count, maximum INR, and maximum aPTT; the clinical-only model included all non-coagulation predictors; and the combined model used the complete common predictor set. All three used the same XGBoost specification. Incremental value was assessed primarily by the paired bootstrap AUROC difference between the combined and clinical-only models, while comparison with the coagulation-only model characterized the standalone prognostic information contained in routine coagulation measurements.

Decision-curve analysis was used to evaluate potential clinical utility in the broader secondary eICU cohort [28, 29]. This analysis was concentrated in the larger external cohort because its greater number of outcome events provided more stable threshold-dependent estimates. Net benefit was compared across candidate risk thresholds, with operating characteristics and bootstrap net-benefit estimates summarized at thresholds of 10%, 20%, and 30%. Decision-curve results were interpreted as exploratory because no specific treatment action or clinical operating threshold was prespecified. Feature contributions for the final XGBoost model were evaluated using additive tree-contribution values generated with pred contribs=True, corresponding to SHAP-style feature attributions [30]. Global importance was summarized using the mean absolute contribution value for each predictor in the final MIMIC-IV development cohort. These values were used to describe model behavior and were not interpreted as causal effects.

Two additional sensitivity analyses assessed the robustness of the primary modeling choices. First, class-weighted models were evaluated using class weight=balanced for logistic regression, class weight=balanced subsample for random forest, and scale pos weight equal to the ratio of non-events to events in the relevant MIMIC-IV fitting data for XGBoost. The GCS-based logistic-regression benchmarks were evaluated under the corresponding weighting strategy. Second, an XGBoost model using native missing-value handling was evaluated without mean imputation. These analyses were not used to modify the primary model specification after external evaluation.

### 2.6 Statistical Analysis and Software

Continuous cohort characteristics were summarized using medians and interquartile ranges, while categorical variables were summarized using counts and percentages. Bootstrap confidence intervals were percentile-based and used 2,000 resamples. External cohorts were used exclusively for validation and were not used for model selection, preprocessing estimation, hyperparameter optimization, or post hoc modification of the primary analysis pipeline.

Analyses were conducted in Python 3.10.11. Data processing used NumPy 2.2.6 and pandas 2.3.2. Logistic regression, random forest, preprocessing pipelines, and cross-validation were implemented using scikit-learn 1.7.2 [31]. XGBoost analyses used XGBoost 3.2.0 [24]. Statistical calculations used SciPy 1.15.3, and figures were generated using Matplotlib 3.10.6.

### Use of generative AI

OpenAI ChatGPT was used during manuscript preparation to assist with manuscript organization, language refinement, and drafting of revisions. All AI-assisted content was critically reviewed and edited by the authors, and all numerical and methodological statements were verified against the original analysis outputs and study materials. The tool was not used to construct study cohorts, generate or modify patient data, perform statistical analyses, train or evaluate the prediction models, or generate the reported figures or results. The authors take full responsibility for the final manuscript.

## 3 Results

### 3.1 Cohort Characteristics and Data Availability

The final MIMIC-IV development cohort included 2,669 patients, of whom 388 died during the indexed ICU stay (14.5%). The primary strict eICU cohort included 396 patients with 39 ICU deaths (9.8%), while the broader secondary eICU cohort included 1,555 patients with 168 ICU deaths (10.8%).

Baseline characteristics differed across the development and external-validation populations (Table 1). Age distributions were broadly similar, with median ages of 68 years in MIMIC-IV, 64 years in the strict eICU cohort, and 68 years in the broader eICU cohort. The clearest difference was neurologic severity. Median minimum GCS was 7 [IQR 5–9] in MIMIC-IV, compared with 13 [7–14] in the strict eICU cohort and 12 [7–14] in the broader cohort. In contrast, distributions of platelet count, INR, aPTT, and several routinely collected physiologic and laboratory variables were broadly comparable across cohorts.

**Table 1.** Baseline characteristics of the MIMIC-IV development and strict and broader eICU validation cohorts. Continuous variables are reported as median [interquartile range] and categorical variables as n (%).

| Characteristic | MIMIC-IV development<br>(n = 2669) | Strict eICU primary<br>(n = 396) | Broader eICU secondary<br>(n = 1555) |
| --- | --- | --- | --- |
| Patients, n | 2669 | 396 | 1555 |
| ICU deaths, n (%) | 388 (14.5) | 39 (9.8) | 168 (10.8) |
| Age, years | 68 [57, 79] | 64 [54, 77] | 68 [56, 79] |
| Female sex, n (%) | 1232 (46.2) | 185 (46.7) | 712 (45.8) |
| Platelet minimum, $\times 10^3/\mu\text{L}$ | 200 [156.8, 251] | 205 [169, 253] | 203 [163.3, 247] |
| INR maximum | 1.2 [1.1, 1.3] | 1.1 [1.0, 1.2] | 1.1 [1.0, 1.3] |
| aPTT maximum, s | 28.7 [26.1, 31.7] | 28.0 [25.7, 30.7] | 28.6 [26.0, 32.0] |
| Heart rate maximum, beats/min | 99 [87, 111] | 103 [93, 116] | 102 [91.3, 117] |
| Heart rate minimum, beats/min | 65 [57, 74] | 64.5 [56, 73] | 64 [56, 73] |
| Respiratory rate maximum, breaths/min | 26 [23, 30] | 26 [23, 32] | 27 [24, 34] |
| Respiratory rate minimum, breaths/min | 12 [10, 14] | 12 [9, 14] | 12 [9, 14] |
| Systolic blood pressure minimum, mmHg | 105 [93, 116] | 104 [90, 115.3] | 100 [86, 113] |
| Temperature minimum, $^{\circ}\text{C}$ | 36.56 [36.39, 36.78] | 36.50 [36.10, 36.78] | 36.50 [36.20, 36.70] |
| Temperature maximum, $^{\circ}\text{C}$ | 37.39 [37.06, 37.89] | 37.30 [37.00, 37.80] | 37.30 [37.00, 37.80] |
| GCS minimum, points | 7 [5, 9] | 13 [7, 14] | 12 [7, 14] |
| WBC maximum, $\times 10^3/\mu\text{L}$ | 10.8 [8.3, 13.9] | 10.8 [8.3, 13.3] | 10.4 [7.9, 13.6] |
| BUN maximum, mg/dL | 17 [12, 23] | 16 [12, 22] | 16 [12, 23] |
| Potassium minimum, mEq/L | 3.8 [3.5, 4.1] | 3.7 [3.4, 4.0] | 3.7 [3.5, 4.1] |
| Potassium maximum, mEq/L | 4.1 [3.8, 4.4] | 3.8 [3.6, 4.2] | 3.9 [3.7, 4.2] |
| Sodium minimum, mEq/L | 139 [136, 141] | 139 [136.3, 141] | 139 [136, 141] |
| Sodium maximum, mEq/L | 141 [138, 143] | 140 [138, 143] | 140 [138, 143] |
| Bicarbonate maximum, mEq/L | 24 [22, 26] | 25 [23, 27] | 25 [23, 27] |
| Total bilirubin maximum, mg/dL | 0.60 [0.40, 0.90] | 0.70 [0.50, 0.90] | 0.70 [0.50, 1.00] |

Abbreviations: aPTT, activated partial thromboplastin time; BUN, blood urea nitrogen; GCS, Glasgow Coma Scale; ICU, intensive care unit; INR, international normalized ratio; WBC, white blood cell count.

Predictor availability also differed between databases (Table 2). Most routinely collected vital signs and chemistry measurements had relatively low missingness, and temperature was missing in approximately 1% or less of both external cohorts while minimum systolic blood pressure was complete. In contrast, missingness was substantially higher in eICU for INR (54.8% in the strict cohort and 51.6% in the broader cohort), aPTT (68.4% and 66.5%), bilirubin (67.4% and 67.7%), and GCS (21.0% and 25.0%).

**Table 2.** Predictor missingness across the development and external-validation cohorts. Values are percentages of patients with missing measurements during the first 24 hours.

| Predictor | MIMIC-IV development | eICU strict primary | eICU broader secondary |
| --- | --- | --- | --- |
| Age | 0.0 | 0.0 | 0.0 |
| Female sex | 0.0 | 0.0 | 0.0 |
| Platelet minimum | 5.4 | 12.9 | 13.2 |
| INR maximum | 14.0 | 54.8 | 51.6 |
| aPTT maximum | 14.8 | 68.4 | 66.5 |
| Heart-rate maximum | 0.1 | 1.5 | 2.1 |
| Heart-rate minimum | 0.1 | 1.5 | 2.1 |
| Respiratory-rate maximum | 0.4 | 5.8 | 6.3 |
| Respiratory-rate minimum | 0.4 | 5.8 | 6.3 |
| Systolic blood pressure minimum | 5.5 | 0.0 | 0.0 |
| Temperature minimum | 0.4 | 0.8 | 1.0 |
| Temperature maximum | 0.4 | 0.8 | 1.0 |
| GCS minimum | 0.1 | 21.0 | 25.0 |
| WBC maximum | 5.4 | 12.9 | 11.9 |
| BUN maximum | 5.0 | 10.9 | 9.8 |
| Potassium minimum | 4.8 | 10.1 | 9.1 |
| Potassium maximum | 4.8 | 10.1 | 9.1 |
| Sodium minimum | 4.6 | 10.6 | 8.9 |
| Sodium maximum | 4.6 | 10.6 | 8.9 |
| Bicarbonate maximum | 4.9 | 11.4 | 11.2 |
| Bilirubin maximum | 54.6 | 67.4 | 67.7 |

### 3.2 Internal Validation

All three multivariable models showed strong discrimination in the MIMIC-IV holdout cohort (Table 3). Logistic regression achieved an AUROC of 0.870 (95% CI 0.841– 0.896), random forest 0.859 (95% CI 0.827–0.889), and XGBoost 0.867 (95% CI 0.836– 0.896). Corresponding AUPRC values were 0.521, 0.510, and 0.511, with Brier scores of 0.093, 0.094, and 0.094, respectively.

**Table 3.** Performance of multivariable prediction models and parsimonious clinical benchmarks in internal and external validation.

| Cohort | Model | AUROC | 95% CI | AUPRC | 95% CI | Brier score |
| --- | --- | --- | --- | --- | --- | --- |
| MIMIC-IV holdout | Logistic regression | 0.870 | 0.841–0.896 | 0.521 | 0.427–0.612 | 0.093 |
|  | Random forest | 0.859 | 0.827–0.889 | 0.510 | 0.419–0.619 | 0.094 |
|  | XGBoost | 0.867 | 0.836–0.896 | 0.511 | 0.424–0.616 | 0.094 |
|  | GCS-only | 0.842 | 0.809–0.872 | 0.384 | 0.314–0.460 | 0.100 |
|  | Age + GCS | 0.852 | 0.820–0.881 | 0.457 | 0.373–0.550 | 0.099 |
| eICU strict primary | Logistic regression | 0.804 | 0.731–0.871 | 0.342 | 0.229–0.510 | 0.080 |
|  | Random forest | 0.839 | 0.780–0.895 | 0.419 | 0.282–0.576 | 0.072 |
|  | XGBoost | 0.817 | 0.744–0.884 | 0.406 | 0.269–0.569 | 0.074 |
|  | GCS-only | 0.780 | 0.690–0.859 | 0.335 | 0.214–0.476 | 0.075 |
|  | Age + GCS | 0.781 | 0.696–0.856 | 0.314 | 0.210–0.469 | 0.077 |
| eICU broader secondary | Logistic regression | 0.826 | 0.797–0.853 | 0.317 | 0.264–0.386 | 0.094 |
|  | Random forest | 0.848 | 0.819–0.874 | 0.420 | 0.350–0.504 | 0.077 |
|  | XGBoost | 0.838 | 0.807–0.868 | 0.416 | 0.345–0.497 | 0.080 |
|  | GCS-only | 0.813 | 0.779–0.843 | 0.326 | 0.267–0.389 | 0.083 |
|  | Age + GCS | 0.815 | 0.781–0.844 | 0.359 | 0.291–0.434 | 0.084 |

Five-fold cross-validation produced similar estimates, with mean AUROCs of 0.861*±*0.011 for logistic regression, 0.868*±*0.022 for random forest, and 0.873*±*0.017 for XGBoost. The GCS-only and age-plus-GCS benchmarks achieved holdout AUROCs of 0.842 and 0.852, respectively. Thus, discrimination was broadly similar across the three multivariable algorithms internally, with modest improvement over the parsimonious clinical benchmarks.

### 3.3 External Validation and Clinical Benchmark Comparison

In the primary strict eICU cohort, all three multivariable models retained useful discrimination, although performance was lower than in the internal MIMIC-IV evaluation. Random forest had the highest numerical AUROC at 0.839 (95% CI 0.780–0.895), followed by XGBoost at 0.817 (95% CI 0.744–0.884) and logistic regression at 0.804 (95% CI 0.731–0.871). Random forest also had the highest AUPRC (0.419) and the lowest Brier score (0.072). However, paired bootstrap comparisons did not clearly distinguish the three multivariable models: the random-forest-minuslogistic-regression AUROC difference was 0.035 (95% CI *−*0.013 to 0.086), and the random-forest-minus-XGBoost difference was 0.022 (95% CI *−*0.024 to 0.068).

The simple clinical benchmarks remained predictive in the strict cohort, with AUROCs of 0.780 for GCS alone and 0.781 for age plus GCS. Random forest exceeded the GCS-only benchmark by 0.059 AUROC (95% CI 0.004–0.124), whereas the corresponding differences for logistic regression and XGBoost were less precisely estimated.

Performance in the broader secondary eICU cohort was similar and more precisely estimated. Random forest again had the highest numerical AUROC at 0.848 (95% CI 0.819–0.874), followed by XGBoost at 0.838 (95% CI 0.807–0.868) and logistic regression at 0.826 (95% CI 0.797–0.853). Corresponding AUPRC values were 0.420, 0.416, and 0.317. Pairwise differences among the multivariable models again remained small: random forest exceeded logistic regression by 0.022 AUROC (95% CI *−*0.002 to 0.047) and XGBoost by 0.009 (95% CI *−*0.011 to 0.031).

The GCS-only and age-plus-GCS benchmarks achieved AUROCs of 0.813 and 0.815 in the broader cohort. Random forest exceeded GCS alone by 0.036 AUROC (95% CI 0.011–0.063), while XGBoost exceeded GCS alone by 0.026 (95% CI 0.005– 0.049). Overall, the external results favored the multivariable models over GCS-based risk representation, but did not provide clear evidence that one multivariable algorithm was uniformly superior to the others.

### 3.4 Coagulation Feature-Group Analysis and Model Interpretation

Routine coagulation measurements showed limited standalone predictive value (Table 4). In the MIMIC-IV holdout cohort, coagulation-only XGBoost achieved an AUROC of 0.598 (95% CI 0.540–0.652), compared with 0.866 (95% CI 0.835–0.896) for the clinical-only model and 0.867 (95% CI 0.836–0.896) for the combined model. The same pattern persisted during external validation. In the strict eICU cohort, coagulation-only XGBoost achieved an AUROC of 0.598 (95% CI 0.501–0.689), whereas clinical-only and combined models achieved AUROCs of 0.803 and 0.817, respectively. The paired AUROC difference for combined minus clinical-only was 0.014 (95% CI *−*0.009 to 0.037). In the broader cohort, coagulation-only AUROC was 0.566 (95% CI 0.522–0.614), compared with 0.835 for clinical-only and 0.838 for combined features. The combined-minus-clinical-only AUROC difference was 0.003 (95% CI *−*0.007 to 0.013). Thus, coagulation variables contributed substantially less discrimination than the broader clinical feature set, and their addition did not produce a clearly supported incremental gain in either external cohort.

**Table 4.**
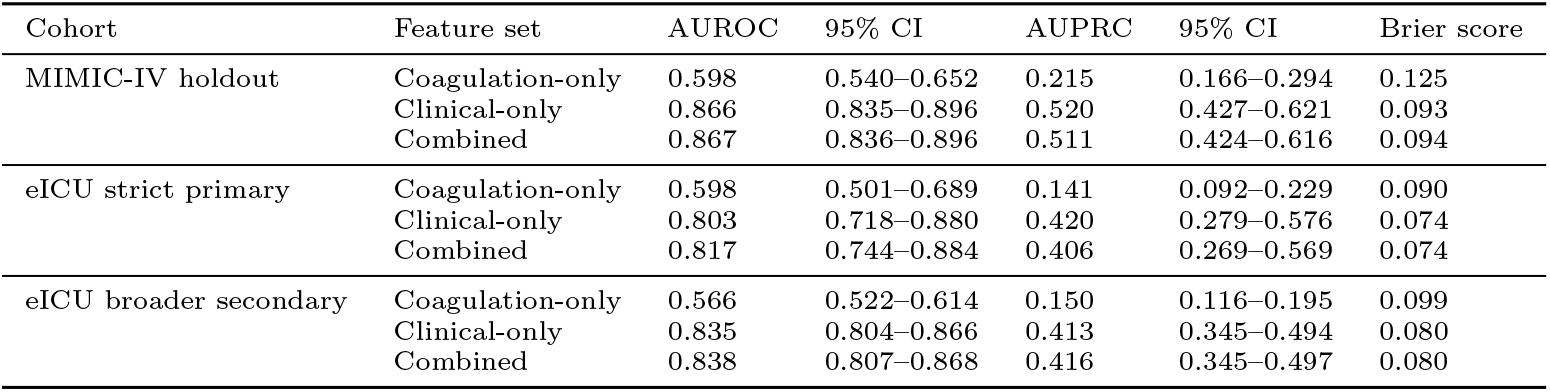
XGBoost feature-group analysis evaluating the predictive contribution of routine coagulation measurements.

| Cohort | Feature set | AUROC | 95% CI | AUPRC | 95% CI | Brier score |
| --- | --- | --- | --- | --- | --- | --- |
| MIMIC-IV holdout | Coagulation-only | 0.598 | 0.540–0.652 | 0.215 | 0.166–0.294 | 0.125 |
|  | Clinical-only | 0.866 | 0.835–0.896 | 0.520 | 0.427–0.621 | 0.093 |
|  | Combined | 0.867 | 0.836–0.896 | 0.511 | 0.424–0.616 | 0.094 |
| eICU strict primary | Coagulation-only | 0.598 | 0.501–0.689 | 0.141 | 0.092–0.229 | 0.090 |
|  | Clinical-only | 0.803 | 0.718–0.880 | 0.420 | 0.279–0.576 | 0.074 |
|  | Combined | 0.817 | 0.744–0.884 | 0.406 | 0.269–0.569 | 0.074 |
| eICU broader secondary | Coagulation-only | 0.566 | 0.522–0.614 | 0.150 | 0.116–0.195 | 0.099 |
|  | Clinical-only | 0.835 | 0.804–0.866 | 0.413 | 0.345–0.494 | 0.080 |
|  | Combined | 0.838 | 0.807–0.868 | 0.416 | 0.345–0.497 | 0.080 |

XGBoost feature-contribution analysis was consistent with these feature-group comparisons (Fig. 1). Minimum GCS was the dominant predictor, with a mean absolute contribution value of 1.309, substantially higher than age (0.319). The next most influential variables were minimum platelet count (0.263), maximum potassium (0.246), maximum BUN (0.237), maximum sodium (0.235), and maximum temperature (0.188). Although platelet count appeared among the higher-ranked individual predictors, the feature-group analysis indicated that routine coagulation measurements as a group contributed little incremental discrimination beyond broader neurologic and systemic information.

**Fig. 1.**
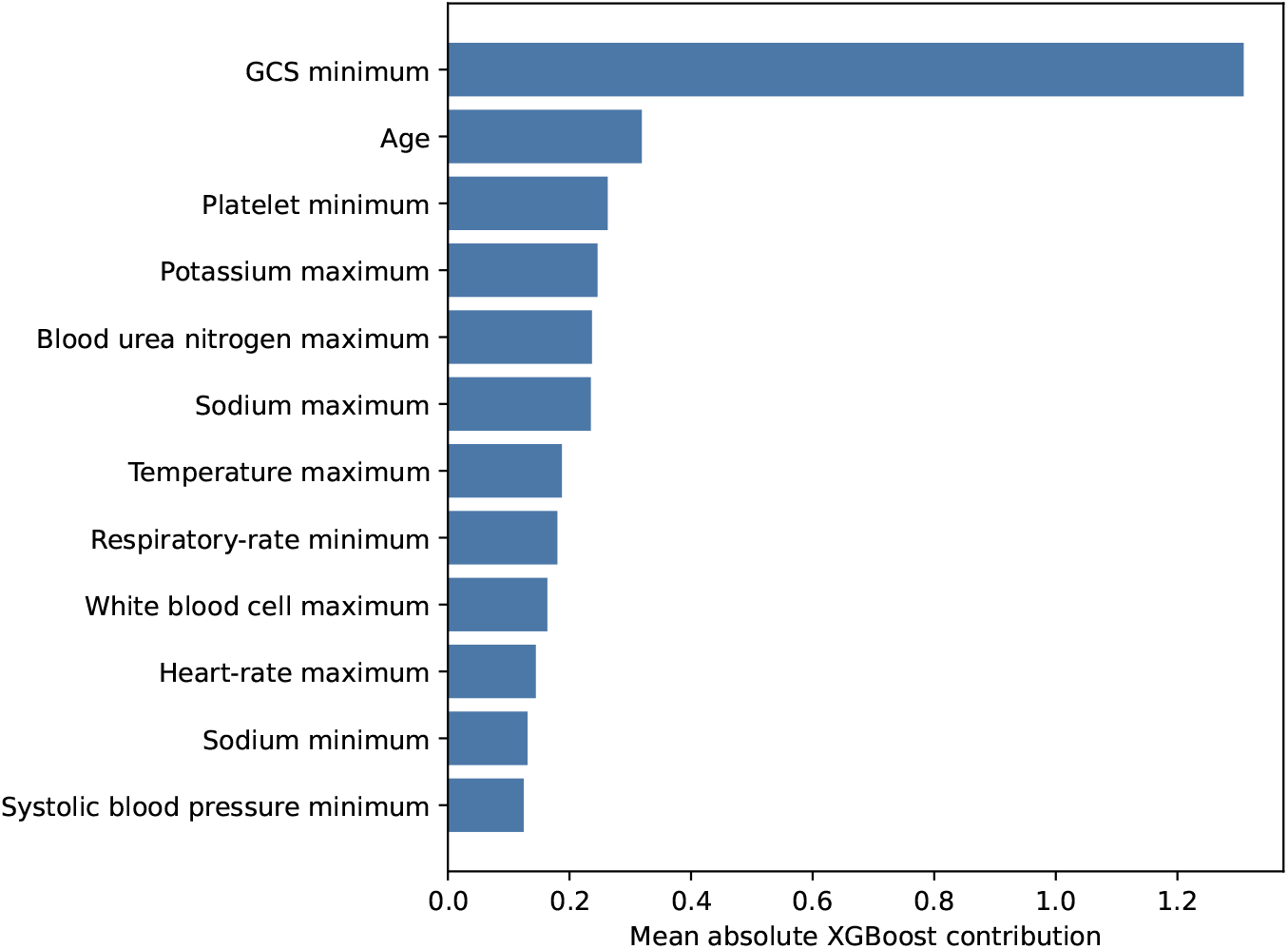
Global feature-contribution ranking for the final XGBoost model in the MIMIC-IV development cohort. Importance is summarized as the mean absolute XGBoost contribution value.

### 3.5 Calibration, Decision-Curve Analysis, and Sensitivity Analyses

Calibration differed across models and external cohorts (Table 5; Figs. 2 and 3). In the strict cohort, the observed ICU mortality rate was 9.8%. Logistic regression predicted a mean risk of 6.3% with a calibration slope of 0.555, while XGBoost predicted 7.7% with a slope of 0.705. Random forest predicted a mean risk of 10.6%, closer to the observed event rate, with a calibration slope of 1.193. Because this cohort contained only 39 deaths, these calibration estimates were relatively imprecise.

**Fig. 2.**
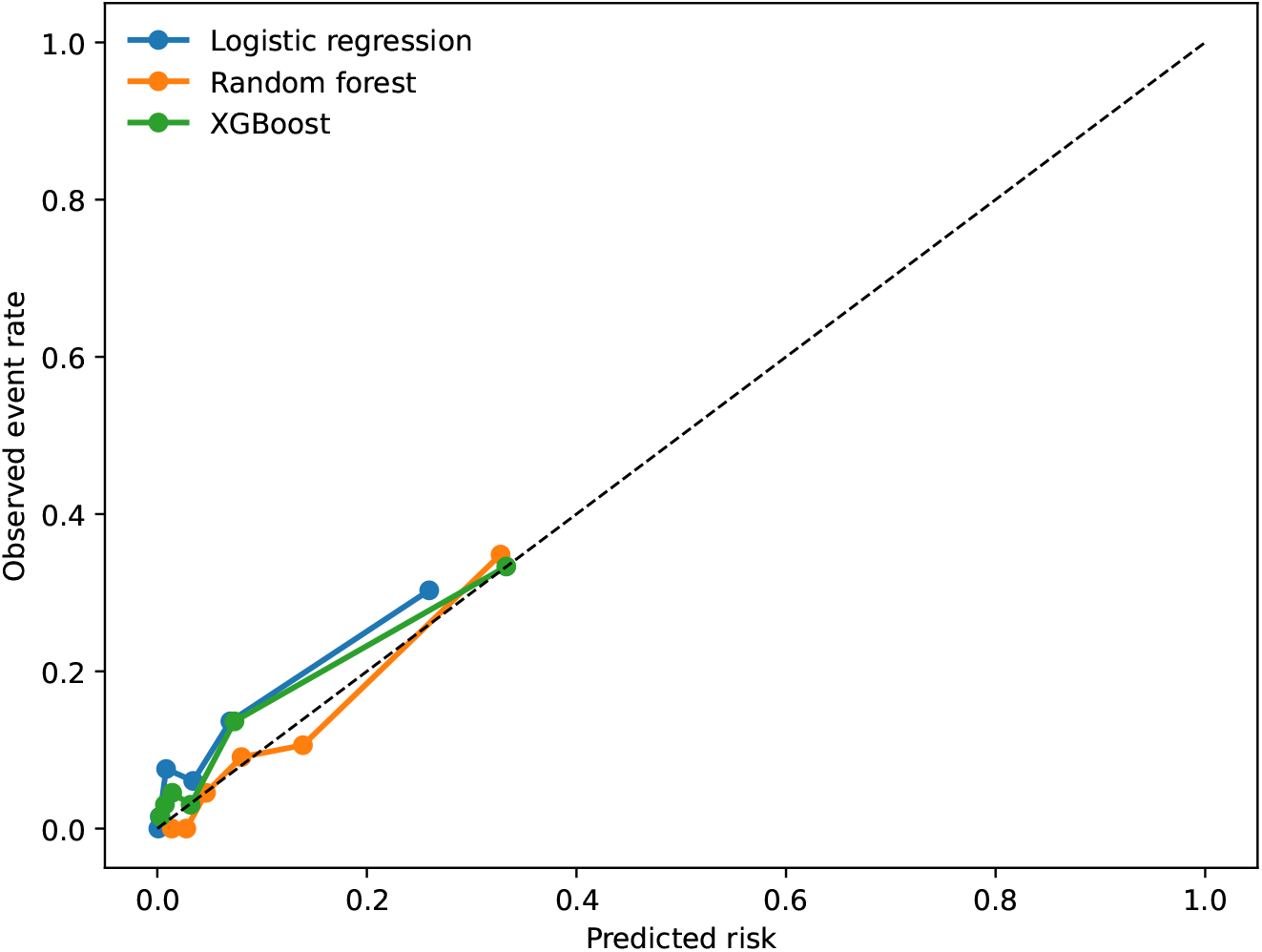
Calibration curves for logistic regression, random forest, and XGBoost in the primary strict eICU cohort. The cohort contained 39 ICU deaths, and calibration estimates should therefore be interpreted cautiously.

**Fig. 3.**
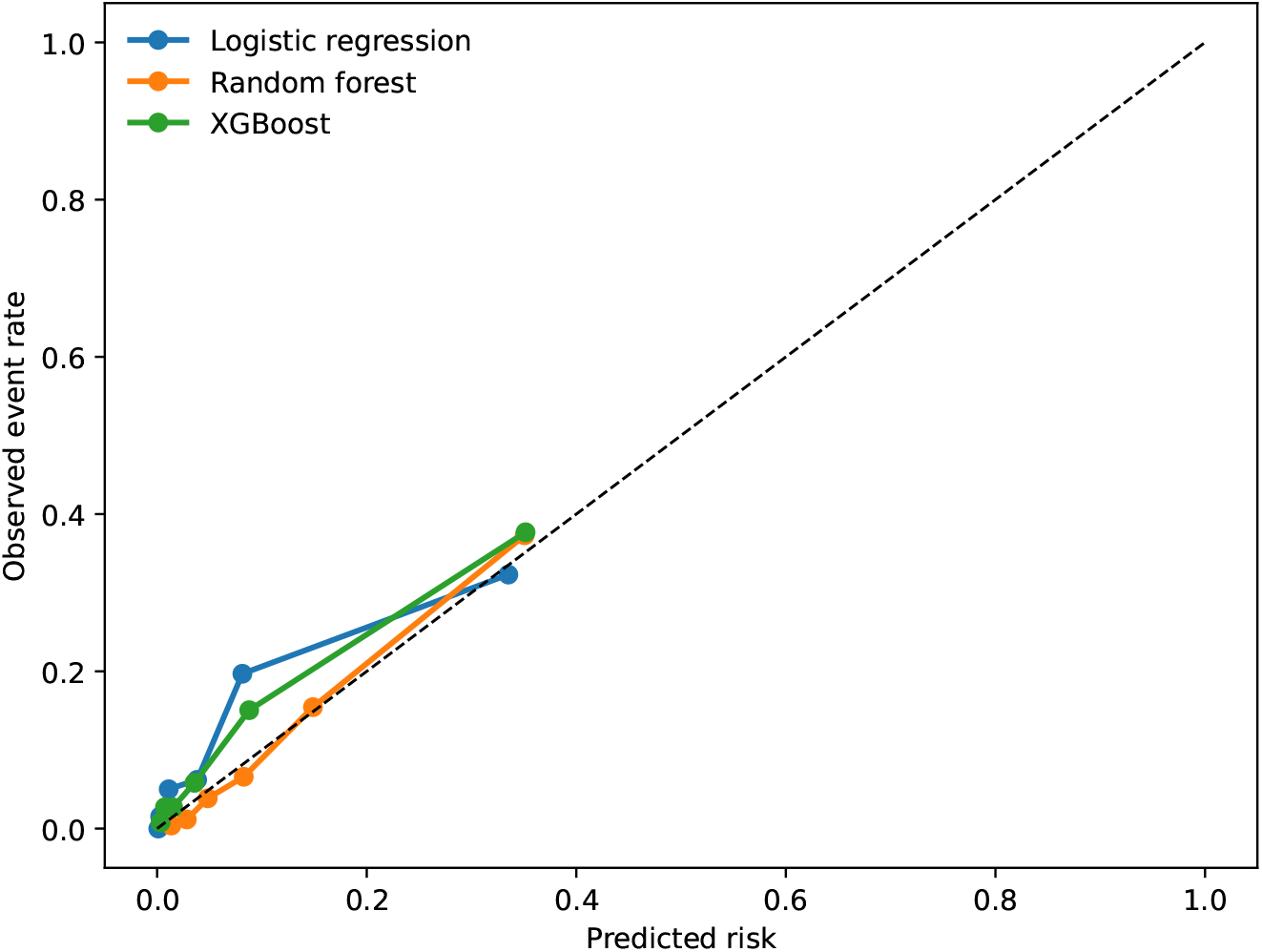
Calibration curves for logistic regression, random forest, and XGBoost in the broader secondary eICU cohort.

**Table 5.** Calibration of the primary multivariable models in the external eICU cohorts.

| External cohort | Model | Calibration intercept | Calibration slope | Mean predicted risk | Observed risk |
| --- | --- | --- | --- | --- | --- |
| Strict primary | Logistic regression | -0.381 | 0.555 | 0.063 | 0.098 |
|  | Random forest | 0.219 | 1.193 | 0.106 | 0.098 |
|  | XGBoost | -0.217 | 0.705 | 0.077 | 0.098 |
| Broader secondary | Logistic regression | -0.617 | 0.482 | 0.078 | 0.108 |
|  | Random forest | 0.267 | 1.205 | 0.112 | 0.108 |
|  | XGBoost | -0.067 | 0.757 | 0.084 | 0.108 |

In the broader cohort, observed ICU mortality was 10.8%. Logistic regression predicted a mean risk of 7.8% and had a calibration slope of 0.482, while XGBoost predicted 8.4% with a slope of 0.757. Random forest again showed the closest agreement in average predicted risk, predicting 11.2%, with a calibration slope of 1.205. Thus, random forest showed the numerically strongest external calibration among the three multivariable models, although none demonstrated uniformly ideal calibration across all measures.

Decision-curve analysis was restricted to the broader secondary cohort (Fig. 4). Random forest and XGBoost generally produced the highest net benefit across the evaluated intermediate risk thresholds. At a 10% risk threshold, net benefit was 0.060 (95% CI 0.045–0.074) for random forest and 0.056 (95% CI 0.043–0.069) for XGBoost, compared with 0.047 for logistic regression and 0.042 for the GCS-only benchmark. At 20%, net benefit remained highest for random forest (0.036, 95% CI 0.023–0.050) and XGBoost (0.034, 95% CI 0.023–0.046). At 30%, corresponding net benefits were 0.020 (95% CI 0.009–0.031) and 0.019 (95% CI 0.009–0.030). These threshold-dependent findings were exploratory because no specific clinical intervention threshold was prespecified.

**Fig. 4.**
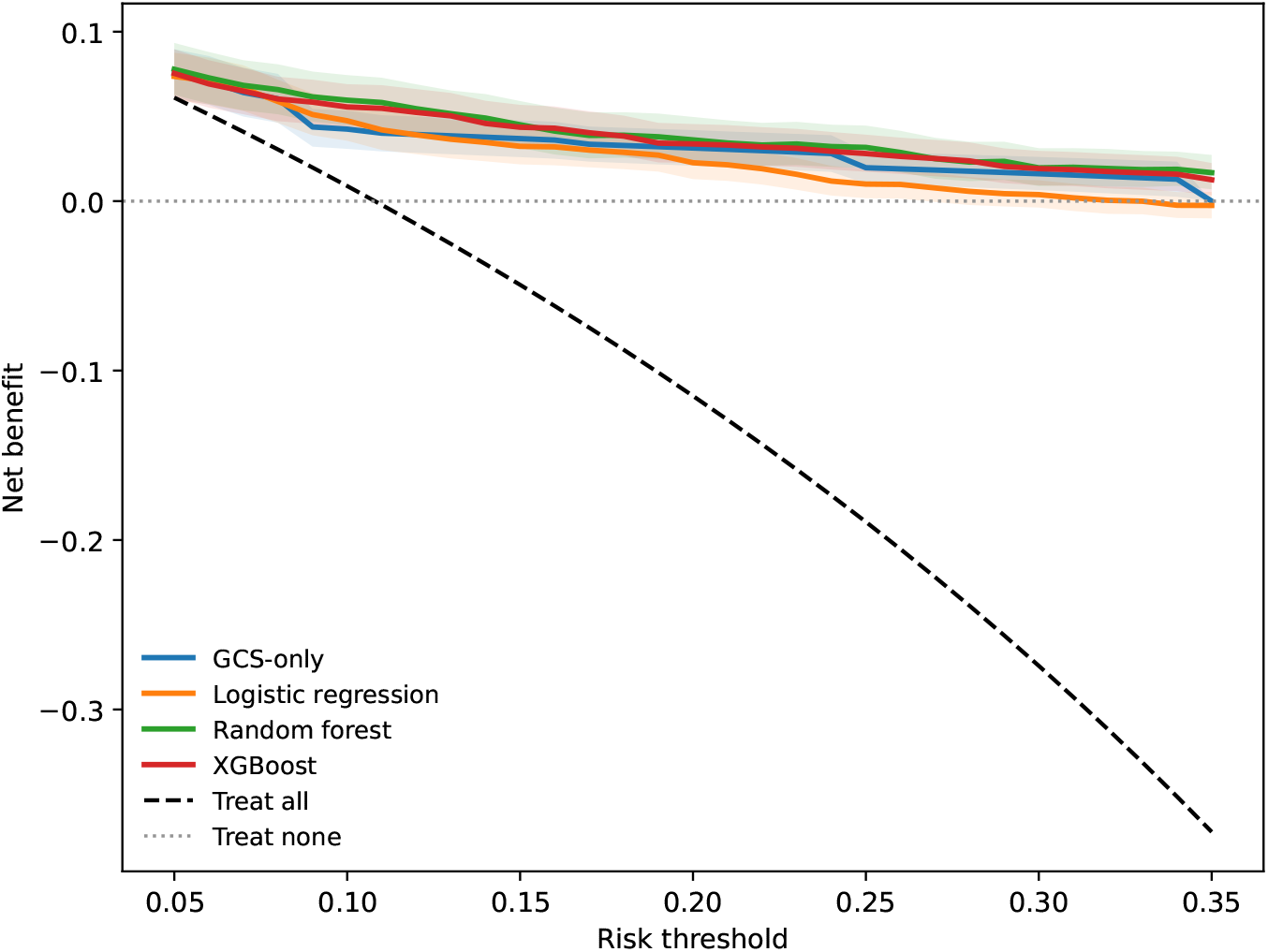
Decision-curve analysis in the broader secondary eICU cohort. Net benefit is shown across predicted-risk thresholds for the GCS-only benchmark and the three multivariable models, together with treat-all and treat-none strategies.

Sensitivity analyses did not materially alter the main findings. Allowing XGBoost to use native missing-value handling rather than mean imputation reduced AUROC from 0.817 to 0.803 in the strict cohort and from 0.838 to 0.828 in the broader cohort; corresponding AUPRC values were 0.386 and 0.387, with Brier scores of 0.076 and 0.082. Class weighting also produced limited changes in discrimination. In the strict cohort, weighted AUROCs were 0.801 for logistic regression, 0.839 for random forest, and 0.803 for XGBoost. In the broader cohort, weighted AUROCs were 0.826, 0.855, and 0.836, respectively. Weighting did not consistently improve probability accuracy and did not change the overall model ranking or the interpretation of the primary unweighted analyses.

## 4 Discussion

### 4.1 Principal Findings

In this cross-database study of ICU mortality after intracerebral hemorrhage, multivariable models developed using information available during the first 24 hours of ICU care retained useful discrimination when transported from MIMIC-IV to eICU. External AUROCs remained approximately 0.80–0.85 in both the high-specificity primary cohort and the substantially larger broader hemorrhagic-stroke/likely-ICH cohort. Random forest showed the numerically strongest external discrimination and calibration, but paired bootstrap comparisons did not provide clear evidence that one multivariable algorithm consistently outperformed the others. This distinction is important: the principal finding is not superiority of a particular machine-learning method, but preservation of useful prognostic information across databases despite differences in case mix, phenotype representation, and predictor availability.

The external populations differed meaningfully from the MIMIC-IV development cohort. In particular, median minimum GCS was 7 in MIMIC-IV but 13 in the strict eICU cohort and 12 in the broader cohort, while ICU mortality declined from 14.5% to 9.8% and 10.8%, respectively. Despite this shift toward less severe neurologic presentations, discrimination remained relatively stable across the two eICU phenotype definitions. The consistency of performance in the strict and broader cohorts suggests that the first-24-hour clinical signal was not dependent on a narrowly defined external sample. At the same time, the observed case-mix differences reinforce that successful transport of discrimination does not imply equivalence of absolute risk across populations.

Simple GCS-based models remained strong predictors externally. GCS alone achieved AUROCs of 0.780 in the strict cohort and 0.813 in the broader cohort, and adding age produced little additional improvement. The multivariable models therefore provided incremental rather than transformative gains over neurologic severity alone. Random forest showed a statistically supported improvement over GCS alone in both external cohorts, while XGBoost also showed a supported improvement in the broader cohort. These findings favor interpreting the multivariable models as complementary risk-stratification tools rather than replacements for established clinical assessment.

Routine coagulation measurements provided a different result. Platelet count, INR, and aPTT alone showed weak discrimination, and adding them to the broader clinical predictor set did not produce a clearly supported improvement in external AUROC. Although platelet count ranked relatively highly in the XGBoost featurecontribution analysis, the feature-group comparison showed that its importance within a fitted model should not be interpreted as evidence that the coagulation domain adds substantial independent predictive information. More broadly, this illustrates why variable-importance rankings and incremental model performance address different questions.

### 4.2 Comparison With Prior Work and Clinical Interpretation

Previous machine-learning studies have demonstrated that mortality after cerebral hemorrhage or spontaneous ICH can be predicted using routinely collected clinical information. Nie et al. reported useful discrimination using multiple machine-learning approaches in critically ill patients with cerebral hemorrhage, while Yap et al. more recently developed high-performing mortality models specifically among ICU patients with spontaneous ICH [10, 11]. Matsumoto et al. further demonstrated the value of integrating clinical and imaging information and directly compared machine-learning approaches with the conventional ICH Score [12]. Together, these studies support the broader premise that nonlinear multivariable models can capture prognostic information beyond individual bedside measurements.

Our findings are consistent with this literature in identifying neurologic severity as the dominant source of predictive information. Minimum GCS was by far the largest contributor to the final XGBoost predictions, and the relatively strong performance of the GCS-only benchmark confirms that much of the mortality signal after ICH is already contained in early neurologic examination. The comparatively modest additional discrimination obtained from broader physiologic and laboratory information is therefore clinically plausible. Importantly, the present models were not designed to replace the ICH Score. Hematoma volume, intraventricular extension, and sufficiently detailed hemorrhage location were not consistently available as structured cross-database predictors, preventing reconstruction of the complete score. The present approach is better understood as a structured electronic-health-record model for risk reassessment at a 24-hour ICU landmark, complementary to rather than substitutive for imaging-based neurologic prognostication.

Recent studies have increasingly addressed model transportability rather than relying solely on random internal validation. Yang et al. used first-day clinical information to predict 30-day mortality after ICH and evaluated their models across MIMIC database versions, while Chen et al. developed time-to-event models in eICU and externally evaluated them in MIMIC-IV [13, 14]. The current study extends this line of work in several respects. First, it addresses a fixed 24-hour landmark and subsequent ICU mortality rather than admission-time or time-to-event prediction. Second, external evaluation was explicitly separated into a high-specificity phenotype aligned with the MIMIC-IV ICD-defined population and a broader phenotype used to test transportability under less restrictive case ascertainment. Third, model performance was interpreted against simple GCS-based benchmarks and with calibration, paired bootstrap comparisons, and decision-curve analysis rather than discrimination alone. The similarity of AUROC across the strict and broader eICU cohorts is particularly relevant to cross-database validation. Clinical databases rarely encode neurologic diagnoses identically. MIMIC-IV permits a relatively concise ICD-based definition, whereas eICU relies heavily on hierarchical and free-text diagnosis descriptions. A highly restrictive external definition may improve disease specificity but reduce event counts and precision, whereas broader definitions increase statistical stability at the potential cost of phenotype heterogeneity. The two-cohort strategy provides complementary information: the strict cohort offers greater confidence in phenotype alignment, while the broader cohort demonstrates that the overall performance pattern persists in a larger and more heterogeneous population. This does not substitute for radiographic adjudication, but it makes the uncertainty introduced by phenotype definition explicit rather than concealing it within a single external sample.

The coagulation findings also refine prior literature. Coagulation abnormalities have established clinical relevance in ICH, and INR-and platelet-based measures have been associated with hospital outcomes after spontaneous hemorrhage [15]. Association with outcome, however, is not equivalent to incremental predictive value. In the present study, coagulation-only models performed poorly, and adding routine coagulation measurements to a model already containing neurologic, physiologic, demographic, and laboratory information produced only minimal changes in AUROC. One interpretation is that these markers capture clinically relevant risk that overlaps substantially with information already represented by broader illness severity. A second consideration is measurement availability: INR and aPTT were missing in more than half of the external population, potentially limiting the transportable information contained in these variables. The results therefore do not imply that coagulation status is clinically unimportant in ICH; rather, they suggest that routinely available platelet count, INR, and aPTT provide limited additional discrimination for this particular prediction task once broader first-day clinical information is available.

Calibration provided a further distinction between predictive ranking and transportable risk estimation. Random forest produced mean predicted risks closest to the observed mortality rate in both external cohorts and calibration slopes near 1.2, whereas logistic regression and XGBoost tended to underestimate average external risk and showed shallower calibration slopes. Such changes are expected when baseline risk, predictor distributions, and predictor–outcome relationships differ between development and deployment populations [17, 27]. Consequently, preservation of AUROC should not be interpreted as evidence that predicted probabilities can be transferred directly into clinical thresholds without local assessment.

Decision-curve analysis in the broader cohort showed numerically greater net benefit for random forest and XGBoost across several intermediate risk thresholds, but these results should remain secondary. No treatment, escalation pathway, or operating threshold was prespecified for this study, and net benefit alone cannot establish that clinical action based on the models would improve patient outcomes. The models are therefore best viewed as candidates for risk stratification and future decision-support evaluation rather than ready-to-deploy treatment triggers.

### 4.3 Strengths, Limitations, and Future Directions

Several features strengthen the present study. The prediction task was anchored to an explicit 24-hour landmark, separating predictor ascertainment from the subsequent outcome period and defining the population to whom predictions would apply. Model evaluation included both a patient-level holdout analysis and stratified crossvalidation internally, followed by independent multicenter external validation without model updating. External phenotype construction explicitly addressed the mismatch between ICD-coded and free-text diagnosis representations, and the use of strict and broader cohorts allowed phenotype specificity and transportability to be examined separately. The analysis also compared multivariable models with simple GCS-based benchmarks, evaluated discrimination and calibration, used paired bootstrap comparisons rather than interpreting small numerical differences alone, and examined both feature-group incremental value and sensitivity to class weighting and missing-data handling.

Several limitations should also be considered. First, this was a retrospective analysis of clinical databases, and ICH status was determined from diagnosis coding or diagnosis text rather than independent review of neuroimaging. In MIMIC-IV, ICH was identified using hospital-level ICD diagnosis codes, which do not provide reliable diagnosis timing relative to ICU admission; therefore, although all predictors were restricted to the first 24 hours of the indexed ICU stay, we could not verify that the coded ICH diagnosis had been formally established before the 24-hour landmark in every case. The strict eICU cohort was designed to maximize phenotype specificity but cannot be considered a radiographically adjudicated spontaneous ICH cohort, while the broader cohort intentionally accepted greater diagnostic heterogeneity.

Second, imaging-derived predictors central to established ICH prognostication— including hematoma volume, intraventricular extension, and detailed hemorrhage location—were not consistently available in the harmonized structured data. A complete ICH Score therefore could not be reconstructed. This limits direct comparison with standard neurologic prognostic tools and may also place an upper bound on the performance achievable using the present feature set.

Third, the 24-hour landmark defines a conditional target population. The models apply only to patients who remain alive and in the ICU beyond the first 24 hours and should not be interpreted as admission-time prognostic models. Patients who died or were discharged before the landmark were outside the prediction target by design. This improves temporal clarity but limits applicability to the earliest phase of ICH care.

Fourth, predictor availability differed substantially between databases. Missingness of INR, aPTT, bilirubin, and GCS was considerably higher in eICU than in MIMIC-IV. Because GCS was missing in 21.0% and 25.0% of the strict and broader eICU cohorts, respectively, the observed cross-database differences in recorded GCS may partly reflect differential documentation or missingness rather than true differences in neurologic case mix alone. Mean imputation provided a consistent transportable preprocessing strategy, and native-missing XGBoost sensitivity analysis did not materially improve performance, but neither approach eliminates potential bias arising from informative measurement, institutional testing patterns, or differences in documentation. The particularly high external missingness of coagulation measurements should also temper conclusions regarding their lack of incremental value.

Fifth, the primary strict external cohort contained only 39 ICU deaths. Although it provided the closest phenotype alignment, this event count limited precision for calibration estimates and small pairwise model differences. The larger secondary cohort partially addressed statistical precision but did so by accepting a broader phenotype definition.

Sixth, ICU mortality is influenced not only by disease severity but also by treatment decisions, discharge practices, and limitations of life-sustaining therapy. The databases did not permit consistent characterization of these processes across cohorts. We also did not evaluate neurologic functional outcome, 30or 90-day mortality, or quality of life, all of which may be more directly relevant to long-term prognosis after ICH.

Seventh, the bootstrap analysis treated patients as independent observations and did not explicitly model clustering of eICU patients within hospitals. Future multicenter studies could use hierarchical or cluster-aware validation to quantify between-center heterogeneity more directly. Finally, feature-contribution values characterize how the fitted XGBoost model used available predictors; they should not be interpreted as causal effects or as evidence that modifying an individual predictor would alter mortality.

Future work should therefore emphasize prospective or temporally separated multicenter validation with imaging-confirmed ICH phenotypes, integration of radiographic severity measures where feasible, and site-specific calibration assessment. Evaluation of functional and longer-term outcomes would broaden clinical relevance beyond ICU survival. If these models are considered for clinical decision support, prospective studies should additionally define the action attached to a predicted risk, assess calibration and operating thresholds in the target institution, and determine whether model-supported decisions improve monitoring, communication, resource use, or patient-centered outcomes.

## 5 Conclusions

At a 24-hour ICU landmark, multivariable models for subsequent ICU mortality after intracerebral hemorrhage retained useful discrimination when transported from MIMIC-IV to eICU cohorts defined using both a high-specificity primary phenotype and a broader secondary hemorrhagic-stroke/likely-ICH phenotype. Random forest showed the numerically strongest external discrimination and calibration, although pairwise comparisons did not establish clear superiority over the other multivariable approaches. Improvements over simple GCS-based benchmarks were modest, emphasizing that early neurologic severity remained the dominant source of prognostic information.

Routine platelet count, INR, and aPTT showed limited standalone predictive value and did not provide clearly supported incremental discrimination once broader neurologic, physiologic, and laboratory information was incorporated. The consistency of model performance across the two external phenotype definitions suggests reasonable transportability of the overall prediction framework, but differences in calibration, predictor availability, and case mix caution against direct clinical use of transported probability estimates. Further evaluation using imaging-confirmed phenotypes, local recalibration, longer-term and functional outcomes, and prospective clinical workflows is needed before these models can support patient-level decision making.

## Data Availability

The MIMIC-IV database (version 3.1) and the eICU Collaborative Research Database (eICU-CRD, version 2.0) analyzed in this study are available to credentialed researchers through PhysioNet at https://physionet.org/content/mimiciv/3.1/ and https://physionet.org/content/eicu-crd/2.0/, respectively. Access requires PhysioNet credentialing, completion of the required human-subjects research training, and acceptance of the applicable data-use agreements. No new patient data were collected, and patient-level source data or derived analytic datasets are not redistributed because access and reuse remain subject to the applicable PhysioNet data-use agreements. Analysis code is available from the corresponding author on reasonable request, subject to the applicable data-use restrictions.

https://physionet.org/content/eicu-crd/2.0/

## Acknowledgements

The authors acknowledge the creators and maintainers of MIMIC-IV, the eICU Collaborative Research Database, and PhysioNet for developing and maintaining these research resources and making them available to the research community.

## Abbreviations

aPTT: Activated partial thromboplastin time
AUPRC: Area under the precision-recall curve
AUROC: Area under the receiver operating characteristic curve
BUN: Blood urea nitrogen
DCA: Decision-curve analysis
eICU-CRD: eICU Collaborative Research Database
GCS: Glasgow Coma Scale
ICH: Intracerebral hemorrhage
ICU: Intensive care unit
INR: International normalized ratio
MIMIC-IV: Medical Information Mart for Intensive Care IV
SHAP: Shapley additive explanations
TRIPOD+AI: Transparent Reporting of a Multivariable Prediction Model for Individual Prognosis or Diagnosis Plus Artificial Intelligence
WBC: White blood cell count

## Declarations

## Ethics approval and consent to participate

This retrospective secondary analysis used de-identified data from MIMIC-IV and the eICU Collaborative Research Database. The collection and sharing of MIMIC-IV were reviewed by the Institutional Review Board of Beth Israel Deaconess Medical Center, which approved the data-sharing initiative and granted a waiver of informed consent. The eICU Collaborative Research Database contains de-identified health data and was accessed through PhysioNet after completion of the required credentialing, human-subjects research training, and data-use agreements. No direct patient contact occurred, and the authors did not have access to directly identifiable patient information.

## Consent for publication

Not applicable.

## Availability of data and materials

The MIMIC-IV database (version 3.1) and the eICU Collaborative Research Database (eICU-CRD, version 2.0) analyzed in this study are available to credentialed researchers through PhysioNet at https://physionet.org/content/mimiciv/3.1/ and https://physionet.org/content/eicu-crd/2.0/, respectively [21, 22]. Access requires PhysioNet credentialing, completion of the required human-subjects research training, and acceptance of the applicable data-use agreements. No new patient data were collected, and patient-level source data or derived analytic datasets are not redistributed because access and reuse remain subject to the applicable PhysioNet data-use agreements. Analysis code supporting cohort construction, preprocessing, model development, validation, and figure generation is available from the corresponding author on reasonable request, subject to the applicable data-use restrictions.

## Authors’ contributions

SP performed data curation, cohort construction and refinement, feature engineering, formal analysis, model development, validation, visualization, and preparation of the original manuscript draft. YS contributed to methodological review, validation of the analytical and reporting framework, interpretation of the results, manuscript restructuring, and writing, review, and editing. JS, MH, and NK contributed to study development, methodological review, interpretation of the findings, and critical revision of the manuscript. KA contributed to clinical interpretation of the findings and critical revision of the manuscript. GP contributed to methodological review and manuscript revision. MP contributed to study conceptualization, methodology, supervision, project administration, interpretation of the results, and critical revision of the manuscript. All authors read and approved the final manuscript.

## Additional file

**Additional file 1.** PDF. **Supplementary Table S1:** Operational definitions of the strict and broader eICU external-validation phenotypes.

