## Additional file 1: Supplementary Table S1 for "Development and Cross-Database External Validation of 24-Hour Landmark Models for ICU Mortality After Intracerebral Hemorrhage: Incremental Value of Routine Coagulation Markers"

Table S1: Operational definitions of the strict and broader eICU external-validation phenotypes.

| **Domain** | **Strict primary phenotype** | **Broader secondary phenotype** |
|---|---|---|
| Diagnosis source | eICU-CRD diagnosis table (`diagnosisstring`); qualifying terminology could occur in any diagnosis row. | eICU-CRD diagnosis table (`diagnosisstring`); a broader hemorrhage-related text screen was used. |
| Primary inclusion logic | Diagnosis text qualified if it contained: (1) explicit intracerebral, intraparenchymal, or parenchymal hemorrhage, hematoma, or bleed terminology; (2) a recognized brain-location term together with explicit hemorrhage, hematoma, or bleed terminology; or (3) "hemorrhagic stroke" or "hemorrhagic CVA" accompanied by hypertensive or intracerebral/intraparenchymal context. | Retained all strict-eligible patients plus additional broader-eligible stays containing explicit hemorrhage-related wording and no competing exclusion diagnosis. |
| Anatomical-location rule | Location terms such as thalamic, cerebellar, pontine, cortical, or brainstem did not qualify independently and required accompanying hemorrhage-related terminology. | No additional location-only rule was used to override the requirement for hemorrhage-related wording. |
| Explicit exclusions | Traumatic cases; subarachnoid hemorrhage; subdural hemorrhage or hematoma; epidural hemorrhage or hematoma; aneurysm or aneurysmal hemorrhage; postoperative or post-craniotomy hemorrhage; CNS mass, mass lesion, tumor, or metastasis context; skull fracture or concussion; and hemorrhagic transformation or conversion of ischemic stroke. | Additional broad-only stays were retained only when no competing exclusion diagnosis was present. |
| Age restriction | Adults only. | Adults only. |
| ICU length of stay | Greater than 1,440 minutes. | Greater than 1,440 minutes. |
| Predictor window | Event offsets 0–1,440 minutes after ICU admission. | Event offsets 0–1,440 minutes after ICU admission. |
| Mortality outcome | ICU death defined by `unitdischargestatus = 'Expired'`. | Same definition. |
| Patient-level selection | First eligible ICU stay per `uniquepid`. | First eligible ICU stay per `uniquepid`, applied after broader phenotype eligibility was determined. |
| Final analysis cohort | 396 patients; 39 ICU deaths. | 1,555 patients; 168 ICU deaths. |
| Relationship between cohorts | All 396 strict-cohort patients were represented in the broader cohort at the patient level. | Of the 396 strict-cohort patients, 393 contributed the identical indexed ICU stay in both analyses; three contributed an earlier broader-eligible ICU stay because first eligible stay was selected separately under the broader phenotype. |

*Abbreviations:* CNS, central nervous system; CVA, cerebrovascular accident; eICU-CRD, eICU Collaborative Research Database; ICU, intensive care unit.
